# A gene-based clustering approach reveals QSOX1/IL1RAP as promising biomarkers for the severity of non-alcoholic fatty liver disease

**DOI:** 10.1101/2023.07.26.23293038

**Authors:** Wenfeng Ma, Jinrong Huang, Benqiang Cai, Mumin Shao, Xuewen Yu, Mikkel Breinholt Kjær, Minling Lv, Xin Zhong, Shaomin Xu, Bolin Zhan, Qun Li, Qi Huang, Mengqing Ma, Lei Cheng, Yonglun Luo, Henning Grønæk, Xiaozhou Zhou, Lin Lin

**Affiliations:** Department of Liver Disease, Shenzhen Traditional Chinese Medicine Hospital, Shenzhen, Guangdong 518033, China; Department of Biomedicine, Aarhus University, Aarhus, Denmark; Steno Diabetes Center Aarhus, Aarhus University Hospital, Aarhus, Denmark; Department of Liver Disease, The Fourth Clinical Medical College of Guangzhou University of Chinese Medicine, Shenzhen, 518033, China; Department of Hepatology and Gastroenterology, Aarhus University Hospital, Aarhus, Denmark; Department of Pathology, Shenzhen Traditional Chinese Medicine Hospital, Shenzhen, Guangdong 518033, China; Department of Pathology, The Fourth Clinical Medical College of Guangzhou University of Chinese Medicine, Shenzhen, 518033, China; Department of Clinical Medicine, Aarhus University, Aarhus, Denmark

**Keywords:** Non-alcoholic fatty liver disease, RNA sequencing data integration, non-invasive biomarker, quiescin sulfhydryl oxidase 1, interleukin-1 receptor accessory protein

## Abstract

**Background and Aims:** Non-alcoholic fatty liver disease (NAFLD) is a progressive liver disease that ranges from simple steatosis to inflammation, fibrosis, and cirrhosis. To address the unmet need for new NAFLD biomarkers, we aimed to identify candidate biomarkers using publicly available RNA sequencing (RNA-seq) and proteomics data.

**Methods:** An approach involving unsupervised gene clustering was performed using homogeneously processed and integrated RNA-seq data of 625 liver specimens to screen for NAFLD biomarkers, in combination with public proteomics data from healthy controls and NAFLD patients. Additionally, we validated the results in the NAFLD and healthy cohorts using enzyme-linked immunosorbent assay (ELISA) of plasma and immunohistochemical staining (IHC) of liver samples.

**Results:** We generated a database (https://dreamapp.biomed.au.dk/NAFLD/) for exploring gene expression changes along NAFLD progression to facilitate the identification of genes and pathways involved in the disease’s progression. Through cross-analysis of the gene and protein clusters, we identified 38 genes as potential biomarkers for NAFLD severity. Up-regulation of Quiescin sulfhydryl oxidase 1 (*QSOX1*) and down-regulation of Interleukin-1 receptor accessory protein (*IL1RAP*) were associated with increasing NAFLD severity in RNA-seq and proteomics data. Particularly, the QSOX1/IL1RAP ratio in plasma demonstrated effectiveness in diagnosing NAFLD, with an area under the receiver operating characteristic (AUROC) of up to 0.95 as quantified by proteomics profiling, and an AUROC of 0.82 with ELISA.

**Conclusions:** We discovered a significant association between the levels of QSOX1 and IL1RAP and NAFLD severity. Furthermore, the QSOX1/IL1RAP ratio shows promise as a non-invasive biomarker for diagnosing NAFLD and assessing its severity.

**Lay Summary:** This study aimed to find non-invasive biomarkers for non-alcoholic fatty liver disease (NAFLD). Researchers utilized a new gene clustering method to analyze RNA-seq data from 625 liver samples. The identified biomarkers were further validated using plasma proteomics profiling, enzyme-linked immunosorbent assay (ELISA), and liver immunohistochemical staining (IHC) in three separate groups of healthy controls and NAFLD patients. The study revealed that the levels of QSOX1 were elevated while IL1RAP levels were reduced with increasing severity of NAFLD. Importantly, the ratio of QSOX1 to IL1RAP expression in plasma showed promise as a non-invasive diagnostic tool for assessing the severity of NAFLD, eliminating the reliance on liver biopsy.

**Graphical abstract:** 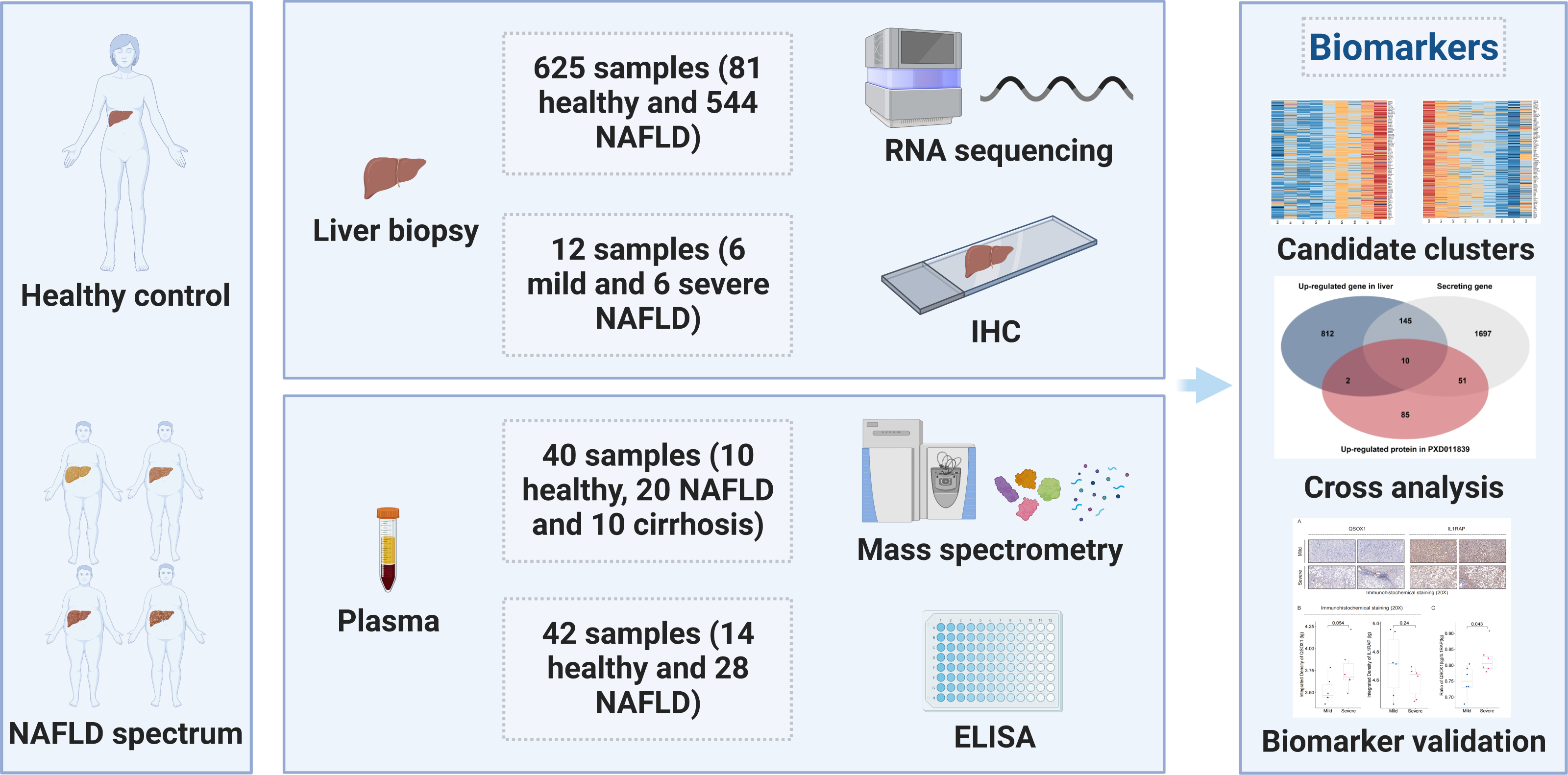

**Highlights:**

- RNA-seq data from 625 liver specimens comprising healthy controls and NAFLD patients with increasing severity were utilized for screening NAFLD biomarkers.
- An unsupervised method for clustering genes based on the similarity of gene expression trajectory across all samples enhanced the discovery of novel effective non-invasive NAFLD biomarkers.
- QSOX1, IL1RAP, and especially the QSOX1/IL1RAP ratio, were found to be associated with NAFLD severity.
- The high sensitivity of the QSOX1/IL1RAP ratio in predicting NAFLD severity was validated with plasma proteomics quantification (AUROC = 0.95) and ELISA (AUROC = 0.82) in two independent patient cohorts.

## Introduction

Non-alcoholic fatty liver disease (NAFLD) is recognized as the hepatic manifestation of the metabolic syndrome, with an estimated global prevalence of around 25-32% (1, 2). The severity of this liver disease ranges from Non-alcoholic Fatty Liver (NAFL) with simple steatosis to Non-alcoholic Steatohepatitis (NASH) with inflammation and fibrosis, which can progress to NASH-induced cirrhosis and increase the risk of hepatocellular carcinoma (HCC).

Liver biopsy is currently the gold standard for histological diagnosis of NAFLD despite its associated side effects such as pain, bleeding, and rare mortality. To address these drawbacks and reduce costs, there is still an unmet need for novel, precise, and cost-effective imaging tools and non-invasive biomarkers (3). Moreover, non-invasive biomarkers are highly needed for replacing repeated liver biopsies when assessing liver histology during pharmacological interventions. Existing NAFLD biomarkers primarily focus on steatosis (e.g., SteatoTest™ or the lipid accumulation product), inflammation (e.g., circulating keratin 18 fragments [CK18), soluble CD163) or fibrosis (e.g., ELF, FibroTest or Pro-C3 tests) (4–8). Despite advancements in biomarker technology, development, and evaluation, an ideal biomarker for the diagnosis, prognosis, and assessment of treatment effects in NAFLD has yet to be identified.

The traditional RNA-seq analysis approach, which relies on established tools such as edgeR (9), DESeq2 (10), and Cufflinks (11), primarily focuses on identifying differentially expressed genes (DEGs) through pairwise comparisons (12). However, for conditions like NAFLD, which involve a complex scoring system and a continuous range of histological variations, this approach has its limitations. NAFLD doesn’t involve transitioning between distinct states but represents a dynamic progression through constant histopathological changes. Pairwise comparisons oversimplify the intricate genetic alterations that occur throughout NAFLD’s development. What’s required is a more advanced analytical method capable of capturing the gradual and overlapping gene expression changes across the entire spectrum of NAFLD. Such an approach would offer a comprehensive representation of NAFLD’s complexity and enhance our understanding of its progression. In recent years, advancements in technologies such as RNA sequencing (RNA-seq), single-cell analysis, and spatial transcriptomics have provided deeper insights into the molecular and cellular processes involved in NAFLD progression (13–15). Large-scale profiling efforts, combined with targeted validation approaches, have led to the discovery of potential biomarkers (16, 17). However, the majority of available RNA-seq data are derived from smaller cohorts of NAFLD patients, which limits the comprehensive understanding of NAFLD severity.

In this study, modularity optimization methods were utilized to cluster genes by employing a graph-based strategy, taking into account the gene expression patterns throughout the progression of NAFLD. We propose that integrating and analyzing these datasets with the unbiased gene-based profiling strategy will provide further insights into the molecular progression of NAFLD and the identification of biomarkers associated with NAFLD severity. In the present study, we identified over 300 NAFLD biomarkers by integrating and analyzing RNA-seq data from 625 liver samples, including their NAFLD activity scores (NAS) and fibrosis scores, along with public proteomics data. We further validated these findings in two independent NAFLD cohorts, demonstrating the potential of the QSOX1/IL1RAP ratio as a non-invasive biomarker for diagnosing NAFLD and assessing its severity.

## Materials and methods

### Data Collection

Genome-wide RNA-seq data of human NAFLD and associated healthy controls were collected from the NCBI GEO (https://www.ncbi.nlm.nih.gov/gds, access date until May 2022). Only datasets that provided detailed NAS and fibrosis scores were included for further investigation, including seven datasets (GSE105127(18), GSE107650(19), GSE126848(20), GSE130970(21), GSE135251(22, 23), GSE162694(24), and GSE167523(25). (**Supplementary Table 1**)

### Data Normalization

The SRA-formatted data were converted into FASTQ format using ‘SraToolkit’ (sratoolkit.2.8.2-1-centos_linux64) (https://github.com/ncbi/sra-tools). Sequencing reads were aligned to the hg19 UCSC RNA sequences Genome Reference Consortium Human Build 37 (GRCh37) using ‘bowtie2’ (bowtie2-2.2.5) (https://rnnh.github.io/bioinfo-notebook/docs/bowtie2.html). Only protein-coding transcripts were considered, and Transcript Per Million (TPM) values were obtained by transforming the mapped transcript reads using ‘RSEM’ (rsem-1.2.12) (https://github.com/deweylab/RSEM). Then, TPM values were then subjected to Trimmed Mean of M-values (TMM) normalization across all samples using ‘metaseqR ’ (metaseqR 1.12.2) (26). The data from various sources involved in this study were integrated and log1p-transformed, followed by batch correction using the ‘removeBatchEffect’ function in the R package ‘limma’ (limma 3.54.2) (https://kasperdanielhansen.github.io/genbioconductor/html/limma.html) (27). Subsequently, the data were expanded (10^x) for further analysis (**Figure 1A**).

**Figure 1.**
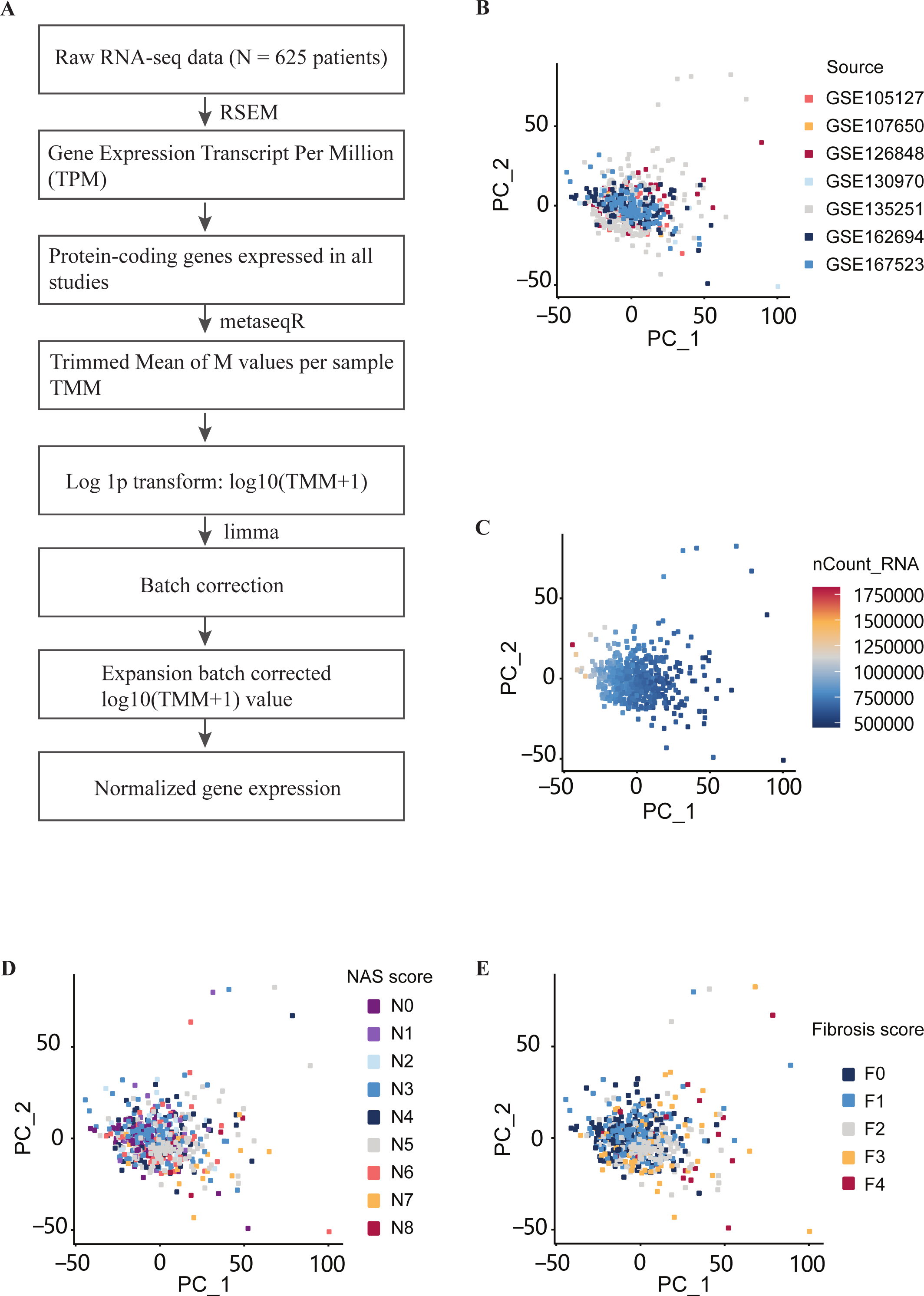
RNA sequencing data processing, integration, and analysis. A. Illustration of data processing. B. Principal component analysis (PCA) based on the origin of datasets. C. PCA based on normalized RNA abundance (nCount). D. PCA based on NAS score. E. PCA based on fibrosis scores.

### RNA-seq Data Analysis and Database Construction

After normalization and batch correction, the RNA-seq data were subjected to Principal Components Analysis (PCA) and unsupervised clustering using the R package ‘Seurat’ (Seurat-4.3.0) (https://satijalab.org/seurat/articles/get_started.html). We utilized the “LogNormalize” method for global-scaling normalization, which normalized the feature expression measurements across different samples for each gene by the total expression. The normalized values were multiplied by a scale factor (default: 10,000) and log1p-transformed. Subsequently, scaling was applied to the identified variable features (default: 2,000). PCA was then performed on the scaled data, with a default setting of computing and storing 50 Principal Components (PCs). To cluster the genes, we employed modularity optimization techniques using a graph-based clustering approach. The dimensions of reduction were set to 1:20, and the resolution parameter was set to 2.3 (28).

To show the gene expression variation during the development of NAFLD associated with both NAS and fibrosis scores, we generated an RNA-seq database using ’ShinyCell’ (https://github.com/SGDDNB/ShinyCell). This database was deployed at https://dreamapp.biomed.au.dk/NAFLD/.

### Proteomics Data Collection and Analysis

The proteomics cohort dataset (PXD011839) includes 10 healthy controls, 10 NAFLD patients with normal glucose tolerance (NAFLD_ngt), 10 NAFLD patients with type 2 diabetes (NAFLD_T2D), and 10 NAFLD patients with cirrhosis (29). We performed the statistical analysis using R-4.3.0 on the dataset (EV1, tab4) (https://www.ncbi.nlm.nih.gov/pmc/articles/PMC6396370/bin/MSB-15-e8793-s003.xlsx). The NAFLD_ngt and NAFLD_T2D groups were merged into a single NAFLD group, resulting in three groups: healthy controls, NAFLD, and cirrhosis.

Similar to RNA-seq analysis, we used ‘Seurat’ (Seurat-4.3.0) and employed the “LogNormalize” method for global-scaling normalization. This method normalized the feature expression measurements across different samples for each protein by the total expression. The normalized values were multiplied by a scale factor (default: 10,000) and log1p-transformed. Subsequently, scaling was applied to the identified variable features (default: 2,000). PCA was performed on the scaled data, with a total of 39 Principal Components (PCs) computed and stored. Additionally, we calculated the log fold-change of the average expression between two groups (avg_log2FC) by comparing the health and NAFLD groups, as well as the NAFLD and cirrhosis groups. By setting avg_log2FC > 0, we selected up- and down-regulated proteins associated with increasing severity of NAFLD.

### Validation of QSOX1 and IL1RAP as Biomarkers in NAFLD

Our objective was to investigate whether *QSOX1* and *IL1RAP* gene expression levels, as well as their encoded proteins, could predict the histological severity of NAFLD. To address this question, we examined the plasma concentrations of QSOX1 and IL1RAP in the proteomics data of healthy and NAFLD cohorts. Additionally, we conducted enzyme-linked immunosorbent assay (ELISA) tests for plasma QSOX1/IL1RAP in a cohort comprising healthy subjects and NAFLD patients recruited at Shenzhen Traditional Chinese Medicine Hospital, China (SZTCMH).

### Human Samples

A total of 28 ultrasound-proven adult NAFLD patients, including NAFLD_ngt and NAFLD_T2D, and 14 healthy controls were enrolled from SZTCMH. Other diagnoses and etiologies, such as excessive alcohol consumption, viral hepatitis, autoimmune liver disease, and the use of steatogenic compounds, were excluded. Archived plasma samples were collected between October and December 2022. Informed consent was obtained from the healthy subjects and NAFLD patients, following the approved clinical protocols of the Ethical Committee of SZTCMH. Clinical information, including body mass index (BMI) and standard biochemistry (liver, kidney, hematology) with metabolic profiling (glucose, insulin, lipids), was collected. Fibroscan with controlled attenuation parameter (CAP) values was performed to assess fibrosis and steatosis. Clinical information for the healthy controls and NAFLD patients can be found in **Supplementary Table 5**.For immunohistochemical staining (IHC), 12 fixed liver tissues were collected from archived histological samples at SZTCMH between 2014 and 2023. These samples were scored based on the NAS score (N0 to N8) and fibrosis score (F0 to F4) by two pathologists (MMS and XWY). Six samples were from mild NAFLD patients (N0-4, F0-2), and six were from severe NAFLD patients (N5-8, F3-4). The clinical study was approved by the Ethical Committee of SZTCMH, and the approved clinical protocols adhere to the Helsinki Declaration (No. K2022-174-01).

### ELISA

Blind ELISA tests were conducted on the collected plasma samples. Randomly assigned sample identifiers and positions were used to ensure blindness to the clinical information and NAFLD stages. The levels of QSOX1 and IL1RAP were measured using QSOX1 ELISA Kits (Catalog No. YJ145587, Lot No. 12/2022 from Enzyme-linked Biotechnology, Shanghai, China) and IL1RAP ELISA Kits (Catalog No. YJ130558, Lot No. 12/2022 from Enzyme-linked Biotechnology, Shanghai, China), respectively. The measurements followed the manufacturer’s instructions and the absorbance values were measured at 450nm. To ensure the reliability of the ELISA Kits, a pre-experiment was conducted three times before the formal experiment.

### Immunohistochemisgtry Assay

We examined the association of QSOX1 and IL1RAP with human NAFLD severity by performing IHC on formalin-fixed and paraffin-embedded liver sections from 6 mild NAFLD patients and 6 severe NAFLD patients. The 3 μm-thick paraffin sections were deparaffinized and rehydrated with distilled water. Antigen retrieval was carried out using pH 9.0 EDTA buffer, followed by 20 minutes of boiling and washing with 1X PBS. Subsequently, the slides were blocked with 1% bovine serum albumin in 1X PBS for 15 minutes and then incubated overnight at 4℃ with QSOX1 (Rabbit anti-human, Catalog No. Ab235444, Lot No. GR3386311-2 from Abcam) or IL1RAP (Rabbit anti-human, Catalog No.35605, Lot No. 4926 from Sabbiotech) antibodies at a concentration of 20 μg/ml. The following day, the slides were washed with 1X PBS and incubated with Goat anti-rabbit IgG H&L (Catalog No. Ab205718, Lot No. ab205718 from Abcam) for 15 minutes at room temperature, followed by another wash with 1X PBS. The images were captured using a light microscope and 3DHISTECH digital scanner (https://www.3dhistech.com/).

The IHC results were analyzed using the software tool ‘**ImagineJ (Fiji)’.** To prevent potential bias, we randomly selected five locations of the same size from each sample at 20x magnification using 3DHISTECH CaseViewer_2.4 (https://www.3dhistech.com/solutions/caseviewer/). Using ‘ImagineJ’, we applied the “Colour Deconvolution” tool with vectors=[H DAB]; followed by selecting the Colour_2 pictures and running “8-bit”. Standard thresholds were used (QSOX1: setThreshold (60, 230), IL1RAP: setThreshold (94, 214)) (30, 31). The average integrated density from the five sites was calculated and used as the integrated density value for each sample, which was then subjected to statistical analysis.

### Statistical analysis

The significance for all statistical tests was two-sided, with P < 0.05. All data analysis was presented in the plots using R-4.3.0, and MedCalc was used to calculate the AUROC, sensitivity, specificity, optimal cutoff value, and sample size.

## Results

### Overview of RNA-seq data and NAFLD patient cohorts

After applying stringent filtering criteria based on the availability of histological NAS and fibrosis scores, five datasets including GSE115193 (32), GSE134422 (33), GSE135448 (34), GSE160016 (35), and GSE164441 (36) were excluded from the analysis, while seven datasets (GSE105127 (18), GSE107650 (19), GSE126848 (20), GSE130970 (21), GSE135251 (22, 23), GSE162694 (24), and GSE167523 (25)) were included. These datasets collectively comprise 81 healthy controls (including healthy obsese individuals) (N0F0) and 544 NAFLD patients. The severity of NAFLD patients was classified based on the NAS score (ranging from N1 to N8) and the fibrosis score (ranging from F0 to F4) using the scoring systems proposed by Brunt (37) and Kleiner (38), respectively (**Table 1, Supplementary Table 1**). A positive correlation was observed between NAS and fibrosis scores (Pearson R = 0.64, P = 4.94E-74, **Table 1**), indicating an association with NAFLD severity.

**Table 1:** Characteristics of the populations studied. *The ratio of gender was estimated according to the gender ratio in the original articles. BMI, body mass index; N, NAS score; F, Fibrosis score.

| **Table 1: Characteristics of the populations studied** |  |  |  |  |  |  |  |
| --- | --- | --- | --- | --- | --- | --- | --- |
| A total of 625 human liver samples of the full histological range from normal, NAFL, NASH to cirrhosis with the NAS (N) and fibrosis (F) scores provided in the database or original articles. |  |  |  |  |  |  |  |
| Sample distribution of NAS and FIB | F0 | F1 | F2 | F3 | F4 | Age (yrs) | BMI (kg/m2) |
| | | | | | | mean ± S.D. | mean ± S.D. |
| N0 (n) | 81 | 1 | 1 | - | 1 | 46.4 ± 10.0 | 35.4 ± 9.1 |
| N1 (n) | 29 | 9 | 1 | - | 1 | 42.8 ± 12.7 | 36.1 ± 6.6 |
| N2 (n) | 27 | 10 | 1 | - | 1 | 51.9 ± 5.8 | 35.1 ± 6.0 |
| N3 (n) | 46 | 72 | 9 | 5 | 3 | 49.8 ± 9.4 | 34.6 ± 8.9 |
| N4 (n) | 30 | 19 | 20 | 15 | 3 | 49.6 ± 9.5 | 33.9 ± 4.6 |
| N5 (n) | 7 | 41 | 75 | 20 | 5 | 52.4 ± 11.7 | 32.5 ± 5.5 |
| N6 (n) | 1 | 18 | 17 | 18 | 3 | 53.2 ± 8.5 | 34.5 ± 5.6 |
| N7 (n) | - | 1 | 14 | 11 | 1 | 49.4 ± 9.8 | 36.3 ± 7.2 |
| N8 (n) | - | - | 2 | 4 | 2 | 54 ± 0.0 | 31.3 ± 0.0 |
| Age (yrs) mean ± S.D. | 46.6 ± 9.6 | 49.0 ± 10.6 | 53.2 ± 11.3 | 54.7 ± 6.0 | 54.8 ± 5.4 | | |
| BMI(kg/m2) mean ± S.D. | 37.1 ± 8.3 | 32.4 ± 5.6 | 33.1 ± 7.0 | 32.5 ± 2.9 | 32.6 ± 2.2 | | |
| Gender n(male/female) | 221(105/116)<br>* | 171<br>(108/63)* | 140<br>(64/76)* | 73<br>(35/38)* | 20 (11/9)* |  |  |

### Normalization and Integration of RNA-seq Data

To address the issue of batch effects resulting from differences in sequencing technology and studies, we processed the integrated data as depicted in **Figure 1A**. Genes with low expression were filtered out, resulting in a total of 17,946 protein-coding genes. Principal component analysis (PCA) demonstrated that normalization effectively eliminated noticeable batch effects (**Figure 1B**). Moreover, neither the NAS nor fibrosis scores appeared to be the main factors contributing to sample separation (**Figure 1D, E**). Instead, the normalized RNA abundance (nCount) in each sample emerged as the key component influencing transcriptome profiles (**Figure 1C**).

### Unsupervised Gene Clustering Identifies Clusters of Genes Associated with NAFLD Severity

To identify genes associated with NAFLD severity, we utilized a previously developed unsupervised gene clustering method based on the similarity of gene expression patterns across each sample (39). We employed gene clustering, grouping genes according to their expression patterns during the progression of NAFLD. By setting a resolution of 2.3, we identified a total of 37 gene clusters (**Supplementary Figure S1**). Notably, cluster 4, consisting of 1021 genes, consistently exhibited increased expression with higher NAS and fibrosis scores (**Figure 2A, Supplementary Figure S2A**). Conversely, cluster 14, comprising 643 genes, showed decreased expression with increasing NAS and fibrosis scores (**Figure 2B**, **Supplementary Figure S2B**). As illustrated in figures (**Figure 2A, B**, **Supplementary Figure S2A, B, C, D**), this approach efficiently clustered genes into distinct groups based on their expression patterns across NAFLD severity stages. It offers a more structured depiction of gene expression variations, enabling a deeper understanding of NAFLD’s molecular pathogenesis. Through visualizing and categorizing these gene expression changes, we can acquire a more comprehensive insight into the underlying mechanisms and factors that propel NAFLD progression.To explore the biological functions of these gene clusters, we performed Gene Ontology (GO) analysis using the R package ‘ClusterProfiler’ (ClusterProfiler-4.8.0). Specifically, we focused on cluster 4, which consisted of up-regulated genes. The GO analysis revealed significant enrichment of genes involved in the fibrosis-related process, such as extracellular matrix (ECM) organization (p.adjust = 9.77E-34), extracellular structure organization (p.adjust = 9.77E-34), external encapsulating structure organization (p.adjust = 1.09E-33), and cell-substrate adhesion (p.adjust = 3.35E-17). Notably, the expression of multiple genes involved in the ECM processes, such as *COL5A3, FBLN5, SPINT2, COL1A1, COL1A2, COL3A1, COL4A1, COL4A4, COL12A1, COL15A1*, and *COL16A1*, showed a gradual up-regulation during the progression of NAFLD (**Figure 2A, Supplementary Figure S3**).

**Figure 2.**
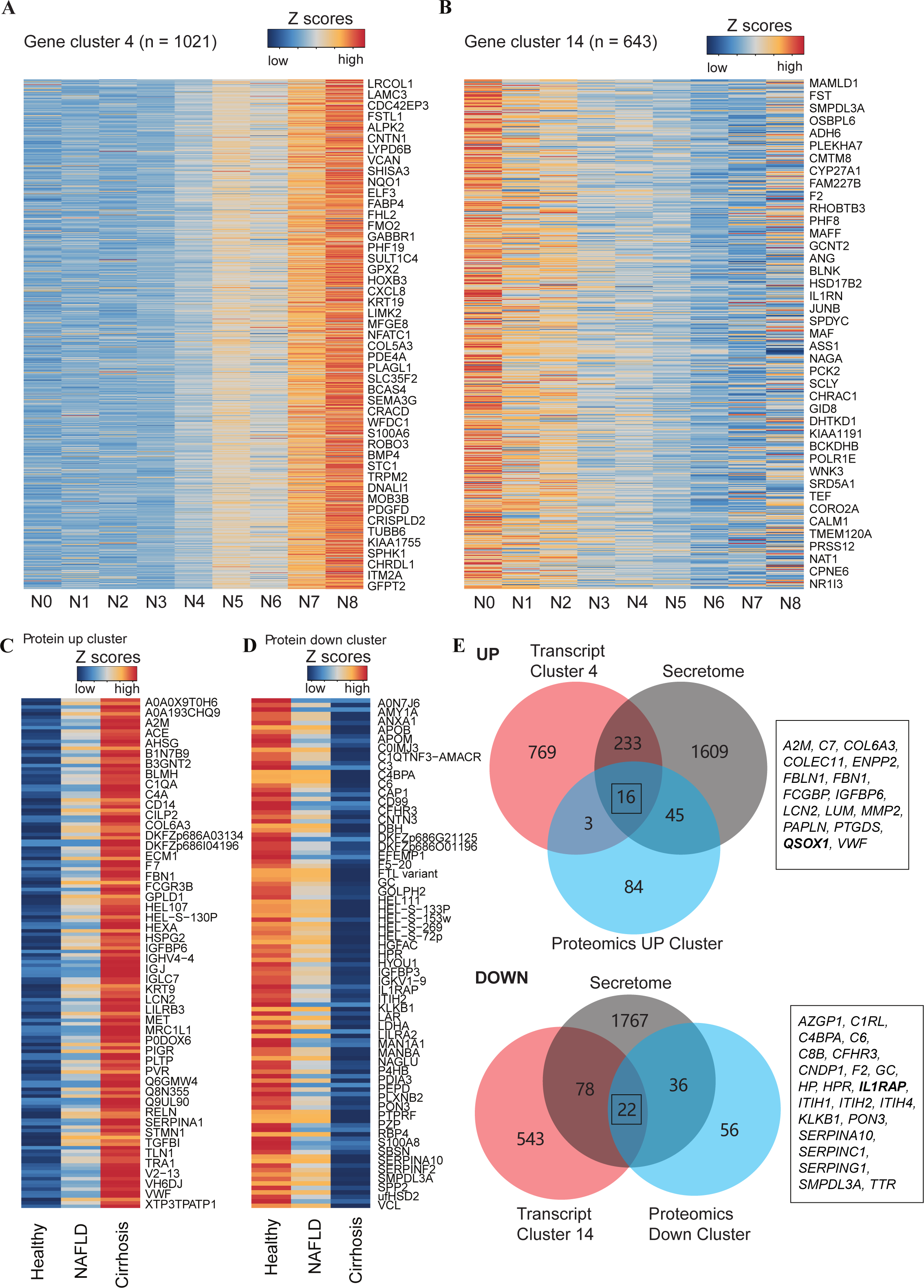
Integrative transcriptome and proteomics analysis to identify NAFLD biomarkers. A. Heatmap presentation of 1021 up-regulated genes in cluster 4 associated with increasing NAS scores. B. Heatmap presentation of 643 down-regulated genes in cluster 14 associated with increasing NAS scores. C. Protein cluster of 148 up-regulated proteins associated with increasing NAFLD severity in PXD011839. D. Protein cluster of 114 down-regulated proteins associated with increasing NAFLD severity in PXD011839. E. (UP) Venn diagram showing 16 overlapping genes between up-regulated genes identified by RNA-seq, up-regulated proteins in the plasma, and secreting proteins. (DOWN) Venn diagram showing 22 overlapping genes between down-regulated genes identified by RNA-seq, down-regulated proteins in the plasma, and secreting proteins.

In contrast, cluster 14, which displayed a reverse correlation with NAS and fibrosis scores, was significantly enriched in metabolic processes, indicating an association between NAFLD progression and attenuated liver metabolism. The down-regulated genes in this cluster were particularly enriched in processes such as organic acid catabolic process (p.adjust = 2.57E-22), carboxylic acid catabolic process (p.adjust = 2.57E-22), small molecule catabolic process (p.adjust = 5.28E-22), alpha-amino acid metabolic process (p.adjust = 6.03E-20), fatty acid metabolic process (p.adjust = 2.60E-10), and alcohol metabolic process (p.adjust = 1.35E-09). Notably, genes encoding enzymes of the Cytochrome P450 superfamily, including *CYP1A2, CYP2C19, CYP2J2, CYP2E1, CYP4A11, CYP4A22, CYP4F11, CYP2C8,* and *CYP3A4*, were down-regulated with increasing NAFLD severity (**Figure 2B**, **Supplementary Figure S4**). For a comprehensive list of all enriched GO terms for genes in cluster 4 and 14, please refer to **Supplementary Table 2**.

In addition, we developed a NAFLD gene expression database (NAFLD-DB) to facilitate the exploration and comparison of all identified protein-coding genes based on NAFLD severity. The NAFLD-DB (https://dreamapp.biomed.au.dk/NAFLD/) was constructed using the ShinyCell framework (40), which was specifically designed for convenient exploration and sharing of single-cell transcriptome data.

### Identification of Candidate Diagnostic Biomarkers

We employed an additional complementary strategy to further refine our list of candidate genes. In this approach, we first analyzed the plasma protein levels from a NAFLD cohort in a proteomics dataset (PXD011839) (29). We selected proteins that showed positive correlations with increasing NAFLD severity (avg_log2FC > 0) and proteins that showed negative correlations. As a result, we identified 148 up-regulated proteins and 114 down-regulated proteins associated with increasing NAFLD severity (**Figure 2C, D**).

The secretome, which consists of secreted proteins, has emerged as a valuable resource for disease diagnostics (41–43). In our study, we aim to identify potential diagnostic markers among the candidate genes, by comparing our gene clusters with the secretome database from the Human Protein Atlas (44). This cross-analysis revealed a total of 349 genes encoding secreted proteins, with 249 genes showing up-regulation and 100 genes showing down-regulation (**Figure 2E**). Notably, our approach successfully identified a comprehensive list of previously known NAFLD diagnostic and prognostic markers, including *ADAMTSL2* (45), *AEBP1* (46), and *BGN* (47) (**Supplementary Table 3**), further validating the effectiveness of our approach.

Next, we intersected the protein-encoding genes of these proteins with the secretome genes and the candidate genes generated from our RNA-seq analysis. Through this cross-comparison, we identified 16 up-regulated secreting genes (*A2M, C7, COL6A3, COLEC11, ENPP2, FBLN1, FBN1, FCGBP, IGFBP6, LCN2, LUM, MMP2, PAPLN, PTGDS, QSOX1, VWF*) and 22 down-regulated secreting genes (*AZGP1, C1RL, C4BPA, C6, C8B, CFHR3, CNDP1, F2, GC, HP, HPR, IL1RAP, ITIH1, ITIH2, ITIH4, KLKB1, PON3, SERPINA10, SERPINC1, SERPING1, SMPDL3A, TTR*) associated with increasing NAFLD severity in both the RNA-seq and proteomics data (**Supplementary Figure S5**).

### QSOX1 and IL1RAP are promising biomarkers for NAFLD severity

To demonstrate the applicability of our NAFLD-DB and validate the association of differential gene expression with increasing NAFLD severity, we selected two representative genes, *QSOX1* and *IL1RAP,* which showed positive and negative correlations with increasing NAFLD severity, and their roles as biomarkers were under explored as compared to other NAFLD biomarkers (**Supplementary Figure S5**). We examined their expression levels in comparison to patients with a NAS or fibrosis score of 0 (N0 or F0). The expression of *QSOX1* was significantly correlated with the severity of NAFLD compared to N0 or F0 patients: N1-4 (p = 0.003), N5-8 (p = 1.9E-10), F1-2 (p = 0.001), F3-4 (p = 6.5E-8) (**Figure 3A, B**). On the other hand, *IL1RAP* expression was significantly lower in patients with increased NAFLD severity compared to N0 or F0: N1-4 (p = 1E-5), N5-8 (p = 4.7E-10), F1-2 (p = 0.00012), F3-4 (p = 0.00013) (**Figure 3C, D**).

**Figure 3.**
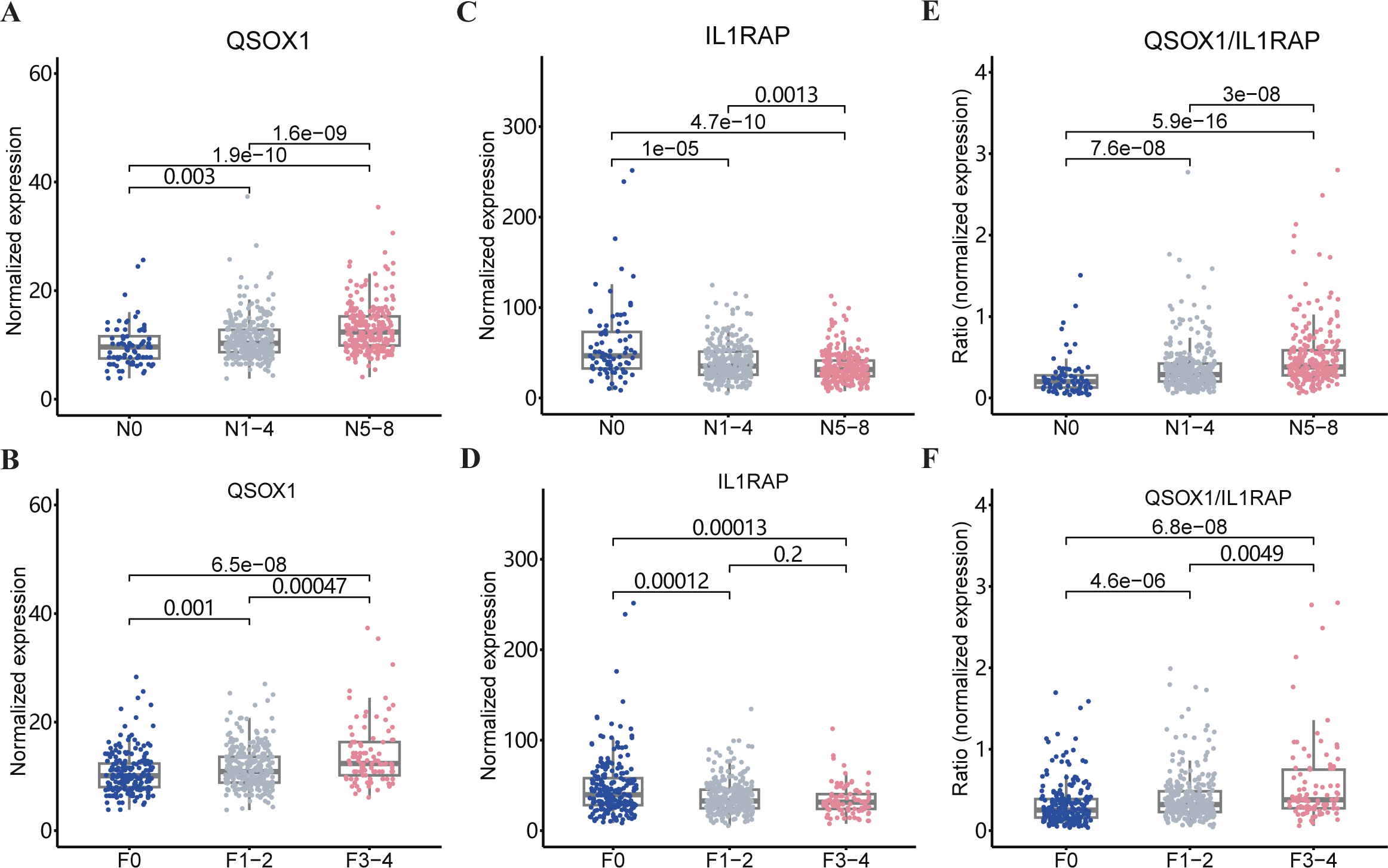
Relationship between QSOX1/IL1RAP and NAS/fibrosis scores in integrative RNA-seq data of the human liver. A. Box plot of QSOX1 gene expressions grouped by NAS scores (N0, N1-4, N5-8). B. Box plot of QSOX1 gene expressions grouped by fibrosis stages (F0, F1-2, F3-4). C. Box plot of IL1RAP gene expressions grouped by NAS scores (N0, N1-4, N5-8). D. Box plot of IL1RAP gene expressions grouped by fibrosis stages (F0, F1-2, F3-4). E. Box plot of QSOX1/IL1RAP gene expression ratio grouped by NAS scores (N0, N1-4, N5-8). F. Box plot of QSOX1/ IL1RAP gene expression ratio grouped by fibrosis stages (F0, F1-2, F3-4). Statistical testing was performed using the Wilcoxon rank sum test, with p-values shown in the plot.

Since *QSOX1* and *IL1RAP* exhibited opposite correlations with NAFLD severity, we further explored whether the ratio of QSOX1/IL1RAP could better distinguish between patient groups. Our results showed that compared to N0 or F0 patients, the ratio of QSOX1 to IL1RAP mRNA levels showed even greater separation: N1-4 (p = 7.6E-8), N5-8 (5.9E-16), F1-2 (4.6E-6), F3-4 (6.8E-8) (**Figure 3E, F**). These findings suggest that the QSOX1/IL1RAP ratio has the potential as a biomarker for diagnosing NAFLD severity.

### Validation of Plasma QSOX1/IL1RAP Levels as Biomarkers for NAFLD Severity with NAFLD Proteomics Cohort

To further validate the potential of QSOX1 and IL1RAP as biomarkers for NAFLD severity, we analyzed the plasma levels of QSOX1 and IL1RAP in a NAFLD proteomics cohort (PXD011839) previously conducted by Niu L and colleagues (29). Consistent with our liver RNA profiling results in livers, the analysis of plasma proteomics data from this independent NAFLD cohort showed a significant increase in plasma QSOX1 levels in patients with NAFLD (Wilcoxon rank sum test, p = 0.021) and cirrhosis (p = 0.049) compared to healthy controls (**Figure 4A**). Conversely, IL1RAP levels were significantly reduced in patients with NAFLD (p = 5.8E-5) and cirrhosis (p = 0.0011) (**Figure 4B**). Moreover, when considering the combined marker of plasma QSOX1/IL1RAP ratio, it demonstrated even greater significance in distinguishing NAFLD (p = 9.3E-6) and cirrhosis (p = 0.00013) patients from the control group, compared to using QSOX1 or IL1RAP alone (**Figure 4C**).

**Figure 4.**
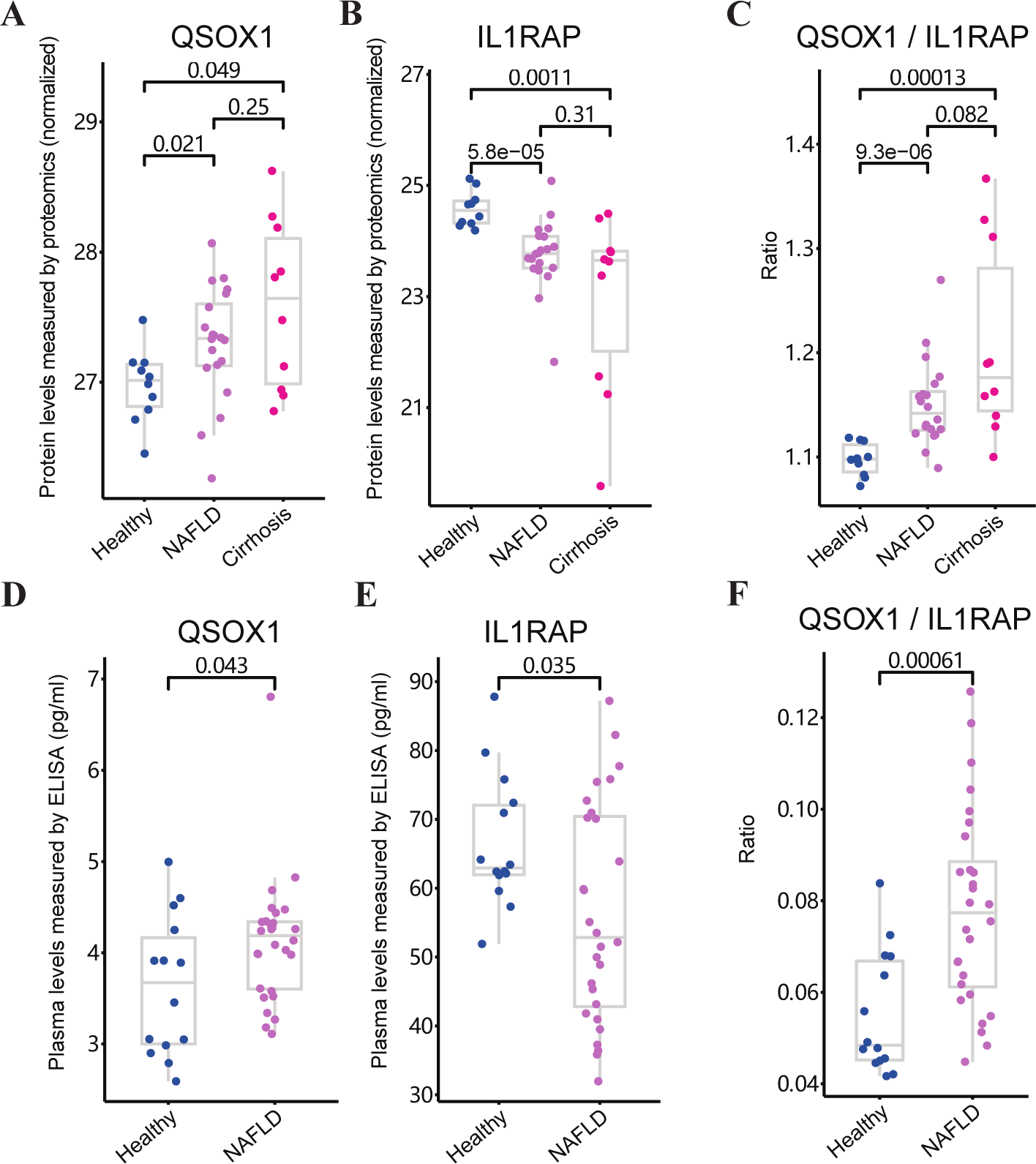
Comparison of plasma protein QSOX1/IL1RAP between healthy individuals and NAFLD at various stages. A-B. Box plots of plasma QSOX1 (A) and IL1RAP (B) protein levels in healthy individuals, NAFLD patients, and cirrhosis patients quantified by mass spectrometry in the NAFLD proteomics cohort. C. Box plots of plasma QSOX1 and IL1RAP protein ratios in healthy controls, NAFLD patients, and cirrhosis patients quantified by mass spectrometry in the NAFLD proteomics cohort. D-E. Box plots of plasma QSOX1 (D) and IL1RAP (E) protein levels in healthy controls and NAFLD groups (pg/ml) measured by ELISA. F. Box plot of plasma QSOX1 and IL1RAP protein ratio. Statistical testing was performed using the Wilcoxon rank sum test, with p-values shown in the plot.

To access the diagnostic sensitivity and specificity of QSOX1, IL1RAP, and their ratio for NAFLD severity, we conducted ROC curve analysis using the ’MedCalc’ tool (30). The sample sizes for each comparison were evaluated and are listed in **Supplementary Table 4**. The AUROC of the QSOX1/IL1RAP ratio for distinguishing NAFLD patients from healthy controls was 0.95, with a cutoff value of 1.12. The sensitivity was determined to be 90%, and the specificity was 100%. Notably, the efficacy of the QSOX1/IL1RAP ratio was superior to that of IL1RAP alone (AUROC=0.92) or QSOX1 alone (not significant). Similarly, when assessing the differentiation between cirrhosis patients and healthy controls, the AUROC of the QSOX1/IL1RAP ratio was 0.96, with a cutoff value of 1.12. The sensitivity was 90%, and the specificity was 100%.

These results indicate that the QSOX1/IL1RAP ratio holds promise as a highly effective biomarker for diagnosing NAFLD severity, surpassing the individual biomarkers alone, and maintaining better sensitivity and specificity in distinguishing NAFLD patients and cirrhosis patients from healthy individuals.

### Validation of QSOX1 and IL1RAP as biomarkers for NAFLD in another patient cohort

To further validate the utility of QSOX1 and IL1RAP as biomarkers for NAFLD, we conducted a validation study in healthy controls and NAFLD patients recruited from the Department of Liver Disease of Shenzhen Traditional Chinese Medicine Hospital. Plasma samples were collected from 14 healthy subjects and 28 newly diagnosed NAFLD patients. Clinical information for the healthy controls and NAFLD patients can be found in **Supplementary Table 5**.

We measured plasma levels of QSOX1 and IL1RAP using an enzyme-linked immunosorbent assay (ELISA). Consistent with our previous findings, plasma levels of QSOX1 (Wilcoxon rank sum test, p = 0.043), IL1RAP (p = 0.035), and the QSOX1/IL1RAP ratio (p = 0.00061) were significantly different between NAFLD patients and controls (**Figure 4D, E, F**).

To assess the diagnostic value of QSOX1 and IL1RAP as non-invasive biomarkers for NAFLD by ELISA, we calculated the AUROC of the QSOX1/IL1RAP ratio in the ELISA test to distinguish NAFLD patients from healthy controls. The QSOX1/IL1RAP ratio exhibited an AUROC of 0.82. Using a cutoff of 0.05, the sensitivity was 93% and the specificity was 57%. In comparison, the AUROC of QSOX1/IL1RAP ratio quantified by ELISA showed less efficacy in distinguishing NAFLD patients from healthy controls (**Supplementary Table 4**), which may be attributed to the sensitivity of protein quantification methods and small sample size.

To further validate the association between QSOX1 and IL1RAP protein levels and NAFLD severity, we assessed their levels in liver biopsies from mild and severe NAFLD patients using IHC. Our results consistently demonstrated a significant correlation between QSOX1 and IL1RAP levels and NAFLD severity (**Figure 5A**). Quantification of QSOX1 and IL1RAP levels based on IHC confirmed that the QSOX1/IL1RAP ratio (p = 0.027) could distinguish the severe NAFLD group (n=6; NAS 5-8, fibrosis score 3-4) from the mild NAFLD group (n=6; NAS 0-4, Fibrosis score 0-2) (**Figure 5B, C**). Collectively, these findings suggest that the QSOX1/IL1RAP ratio holds promise as an effective biomarker for the early diagnosis and prediction of NAFLD severity.

**Figure 5.**
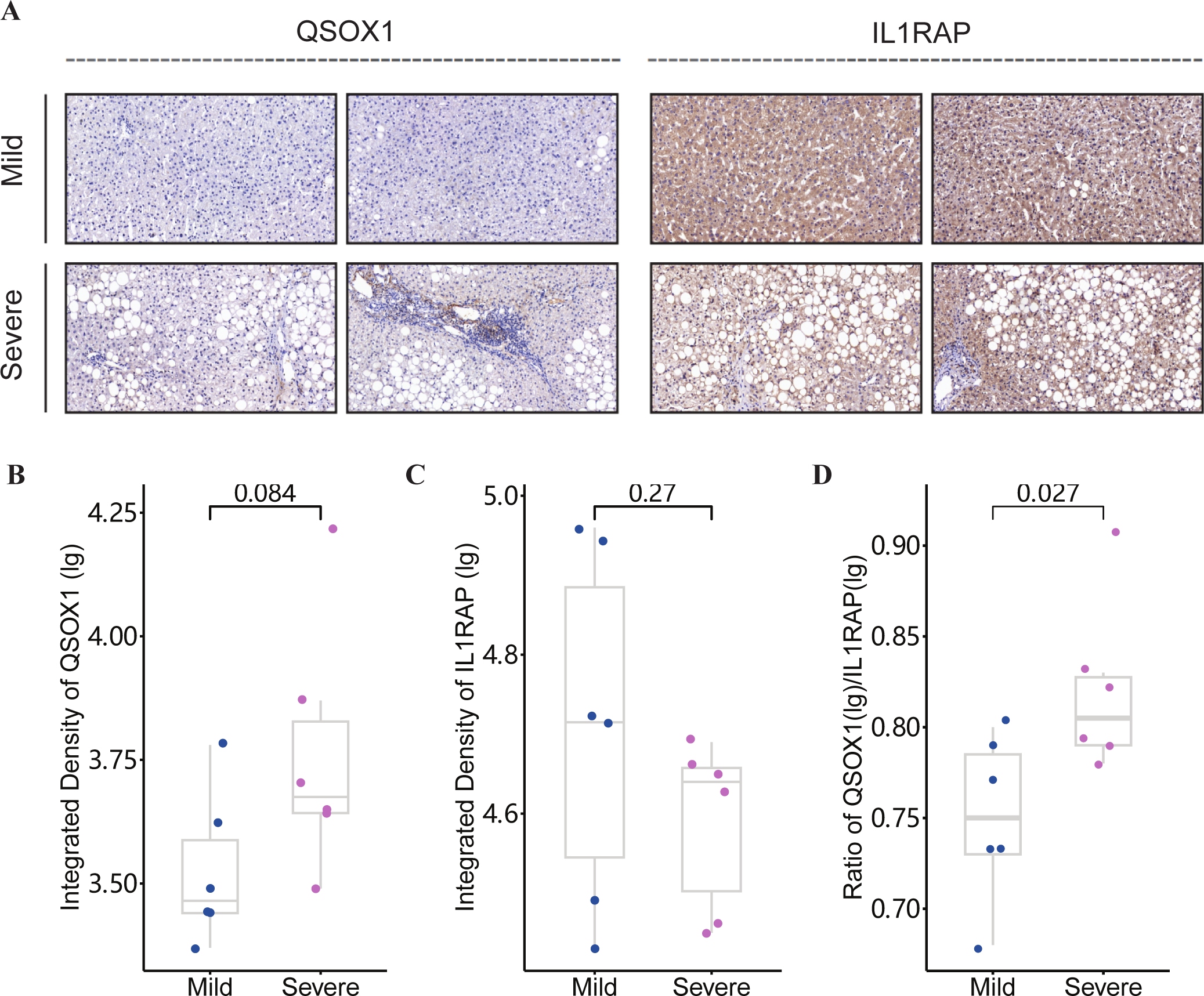
Quantification of liver QSOX1 and IL1RAP levels in NAFLD patients by IHC. A. Representative IHC images of QSOX1 and IL1RAP in liver biopsies from mild NAFLD patients (N0-4, F0-2) and severe NAFLD patients (N5-8, F3-4). B. The integrated density of QSOX1 IHC. Box plots showing the log10 value. C. The integrated density of IL1RAP IHC. Box plots showing the log10 value. D. Box plot of QSOX1 and IL1RAP ratio of the integrated density quantified with IHC. Statistical testing was performed using t-test, with p-values shown in the plot.

## Discussion

This study is the first to integrate publicly available RNA-seq datasets from over 600 NAFLD patients with varying stages of disease severity, combined with proteomics data analysis of publicly available datasets. The key findings suggest that the QSOX1/IL1RAP, and particularly the QSOX1/IL1RAP ratio hold promise as potential biomarkers for NAFLD severity assessment. These results align with recent research highlighting the importance of different transcriptional profiles specific to NAS and fibrosis scores, offering valuable insights into the molecular mechanisms driving disease progression from simple steatosis to inflammation and fibrosis (21, 24).

### The Advantages of Utilizing Integrated RNA-seq Data for Investigating NAFLD Biomarkers

Despite the growing availability of RNA-seq data in this field, many original studies have been limited by small sample sizes and biased sample distribution, making it challenging to accurately decipher transcriptional differences across various stages of NAFLD(18, 20–22, 24). Several studies have attempted to identify diagnostic biomarkers and potential drug targets. For instance, Brosch et al. conducted a positional analysis of transcriptomes across three micro-dissected liver zones from 19 NAFLD patients (18). Suppli et al. demonstrated that immunohistochemical markers offer greater objectivity in distinguishing hepatocyte injury between NASH and NAFL (20). In the pursuit of diagnostic genes and novel drug targets, Hoang et al. studied 6 histologically normal and 72 NAFLD patients, while Pantano et al. studied 31 histologically normal and 112 NAFLD patients. These studies revealed that specific cells proportion and candidate gene signatures can accurately predict fibrosis stage and disease progression (21, 24). Likewise, Govaere et al. observed the correlation between gene expression and histology in a cohort of 10 controls and 206 NAFLD patients (22). In contrast to the studies above that identified sets of potential biomarker genes, Kozumi et al. validated thrombospondin 2 (THBS2) as a noninvasive biomarker for NAFLD. They confirmed its potential in identifying the disease stages among 98 NAFLD patients, and the serum levels of its encoded protein TSP-2, measured by ELISA, showed an AUROC of 0.78 in the diagnosing of NASH among 213 patients with biopsy-proven NAFLD (25). The major challenge of combining and analyzing these diverse datasets lies in achieving homogeneous processing, which requires substantial time and computational resources (48, 49). To generate more robust and compelling results, we employed unbiased integration of comprehensive NAFLD data to profile the liver transcriptome across a broad spectrum of NAFLD severity in our study, incorporating all the aforementioned samples.

### The Superiority of QSOX1 and IL1RAP as Potential Biomarkers of NAFLD

The high prevalence and associated risks of NAFLD have driven global efforts to identify improved diagnostic biomarkers. However, most existing biomarkers are primarily suited for evaluating fibrosis (3, 50–52). The Fibrosis-4 (FIB-4) test commonly used in clinical practice, is sub-optimal for screening purposes, as it carries the risks of both overdiagnosis and false negatives, particularly in patients at risk of chronic liver disease (8). Although the patented ELF™ test was highly recommended for ruling out advanced fibrosis, it comes with higher costs. Several steatosis scores, such as the SteatoTest™ and the fatty liver index (FLI), have been proposed for steatosis detection, but they do not provide substantial additional information beyond routine clinical, laboratory, and imaging examinations conducted in patients suspected of having NAFLD(8). Non-coding RNAs (ncRNAs), which exhibit aberrant expression associated with NAFLD, have emerged as potential biomarkers for NAFLD pathology, and circulating ncRNAs including miR-122 and lncRNAs are proposed as potential biomarkers for NAFLD severity and progression (53–59). Despite the development of new biomarkers, there is still uncertainty surrounding their predictive value, underscoring the urgent need to develop novel, cost-effective, and efficient biomarkers with high sensitivity and specificity for NAFLD prediction and monitoring (4, 60).The approach by Hoang et al. (21) centered on identifying genes with diverse expressions associated with NAFLD severity, inspired us to develop our gene clustering method. Our approach surpasses the constraints of conventional RNA-seq data analysis, which predominantly relies on pairwise comparisons. Instead, it classifies genes according to their dynamic expression patterns, enabling a more comprehensive and dynamic perspective of molecular alterations as NAFLD progresses. This method has the potential to map NAFLD severity and progression solely through gene expressions, thus avoiding invasive procedures like liver biopsies. Moreover, the gene-based scoring system can forecast NAFLD progression, facilitating early interventions for patients at risk of advancing to severe disease stages.Previous studies have explored the relationship between QSOX1, IL1RAP, and NAFLD or steatosis(16, 61). QSOX1 has been suggested as a potential diagnostic biomarker for NAFLD, playing a significant role in lipid metabolism as an enzyme expressed in various tissues, particularly in quiescent fibroblasts (18, 62, 63). IL1RAP is localized in vesicles and cytosol, and it is secreted into the bloodstream. Notably, IL1RAP expression at the RNA level was specifically detected in the liver and hepatocytes (44). Hence, the combination of QSOX1 and IL1RAP as secretome genes and proteins was selected as a potential biomarker combination.

The potential of QSOX1, IL1RAP, and their ratio as biomarkers for NAFLD was demonstrated through the analysis of public RNA-seq and proteomics data, ELISA tests conducted on patients’ plasma, and IHC performed on fixed liver slides. These findings suggest that QSOX1, IL1RAP, and their ratio hold promise as effective biomarkers for NAFLD. Notably, the higher AUROC values for NAFLD diagnosis achieved by QSOX1, IL1RAP, and their ratio highlight their efficacy as NAFLD biomarkers.

### Limitation and Future Prospects

The current study possesses several strengths, including the integration and processing of RNA-seq data from over 600 NAFLD patients with varying degrees of NAFLD severity, as well as validation using proteomics data and samples from NAFLD patients and controls. Furthermore, the well-established database with a user-friendly interface could benefit the research community in exploring differentially expressed genes in NAFLD at various stages. However, there are also limitations to consider. For instance, some samples in the GSE126848 and GSE167523 datasets lacked individual NAS and fibrosis scores. To address this issue, we standardized scores based on their categories in the original articles, and the impact on the results was deemed negligible due to the provision of general stages and unsupervised gene clustering. Machine learning, an essential tool for biomarker validation and sample classification validation, should be employed to train large cohorts of biopsy-proven NAFLD patients and healthy controls. However, this would require an extended recruiting period(64) to determine the sensitivity and specificity of the QSOX1/IL1RAP ratio for NAFLD diagnosis and staging.

Although newer technologies such as single-cell RNA sequencing (scRNA-seq) and spatial sequencing have gained popularity, RNA-seq still serves as a valuable tool in uncovering the pathogenesis of NAFLD (17). Computational analysis limitations make it impractical for large cohort research, and single-cell suspension processing may affect cell abundance and cell type representation, particularly in hepatic ballooning cells in NAFLD (65). Single-nuclei RNA sequencing (snRNA-seq) captures cell frequency more accurately than scRNA-seq but captures lower gene expression. Spatial transcriptomics and proteomics have limitations for discovering invasive biomarkers of NAFLD as they focus on small sampling areas (15). The combination of all these biological tools holds potential for future research.In conclusion, through a novel approach of unsupervised gene clustering performed on integrated RNA-seq data, we have discovered a significant association between QSOX1 and IL1RAP levels and NAFLD severity, with their ratio showing potential as a non-invasive biomarker for diagnosing and assessing the severity of NAFLD. Validation of our plasma-level findings in larger cohorts of liver biopsies is required, but it holds promise as a new tool to diagnose NAFLD severity and reduce the need for liver biopsies. Our approach may lead to the discovery of more NAFLD biomarkers, and the ratios of other up-regulated and down-regulated genes associated with increasing NAFLD severity also have the potential to be verified as potential biomarkers

## Data Availability

The data and code that support the findings of this study are available at https://github.com/cynthia139/NAFLD-RNA-seq.

https://dreamapp.biomed.au.dk/NAFLD/

## Abbreviations

AUROC: area under the receiver operating characteristic
avg_log2FC: log fold-change of the average expression between the two groups
BMI,: body mass index
CK18: circulating keratin 18 fragments
ECM: extracellular matrix
ELISA: enzyme-linked immunosorbent assay
F: Fibrosis score
FIB-4: Fibrosis-4
GEO: Gene Expression Omnibus
GO: Gene Ontology
GRCh37: Genome Reference Consortium Human Build 37
HCC: hepatocellular carcinoma
IHC: immunohistochemistry staining
IL1RAP: Interleukin-1 receptor accessory protein
lg, log10: PCA, Principal components analysis
QSOX1: Quiescin sulfhydryl oxidase 1
RNA-seq: RNA sequencing
N: NAS score
NAFL: Non-alcoholic Fatty Liver
NAFLD: Non-alcoholic fatty liver disease
NAFLD-DB: NAFLD gene expression database
NAFLD_ngt: NAFLD with normal glucose tolerance
NAFLD_T2D: NAFLD with type 2 diabetes
NAS: NAFLD activity scores
NASH: Non-alcoholic Steatohepatitis
ncRNAs: non-coding RNAs
scRNA-seq: single-cell RNA sequencing
snRNA-seq: Single-nuclei RNA sequencing
SZTCMH: Shenzhen Traditional Chinese Medicine Hospital, China
THBS2: thrombospondin 2
TMM: Trimmed Mean of M-values
TPM: Transcript Per Million

## Authors’ contributions

YLL, HG, WFM and LL designed the study and interpreted the data. The analysis strategy has been developed by LL and WFM. WFM, JRH, BQC, MLL, XZ, SMX and MBK collected and assembled the data. WFM drafted the manuscript. WFM, JRH and LC performed data analysis and/or interpretation. Technical support: YLL, LL, JRH, ZXZ, MMS and XWY. Study participant inclusion: ZXZ, WFM, BQC, MLL, XZ, SMX, BLZ, QL, QH, MQM. All authors reviewed and approved the final version of the manuscript.

## Data and code availability statement

The data that support the findings of this study are available from the corresponding author upon reasonable request.

## Supplementary File

**Supplementary Figure S1.**
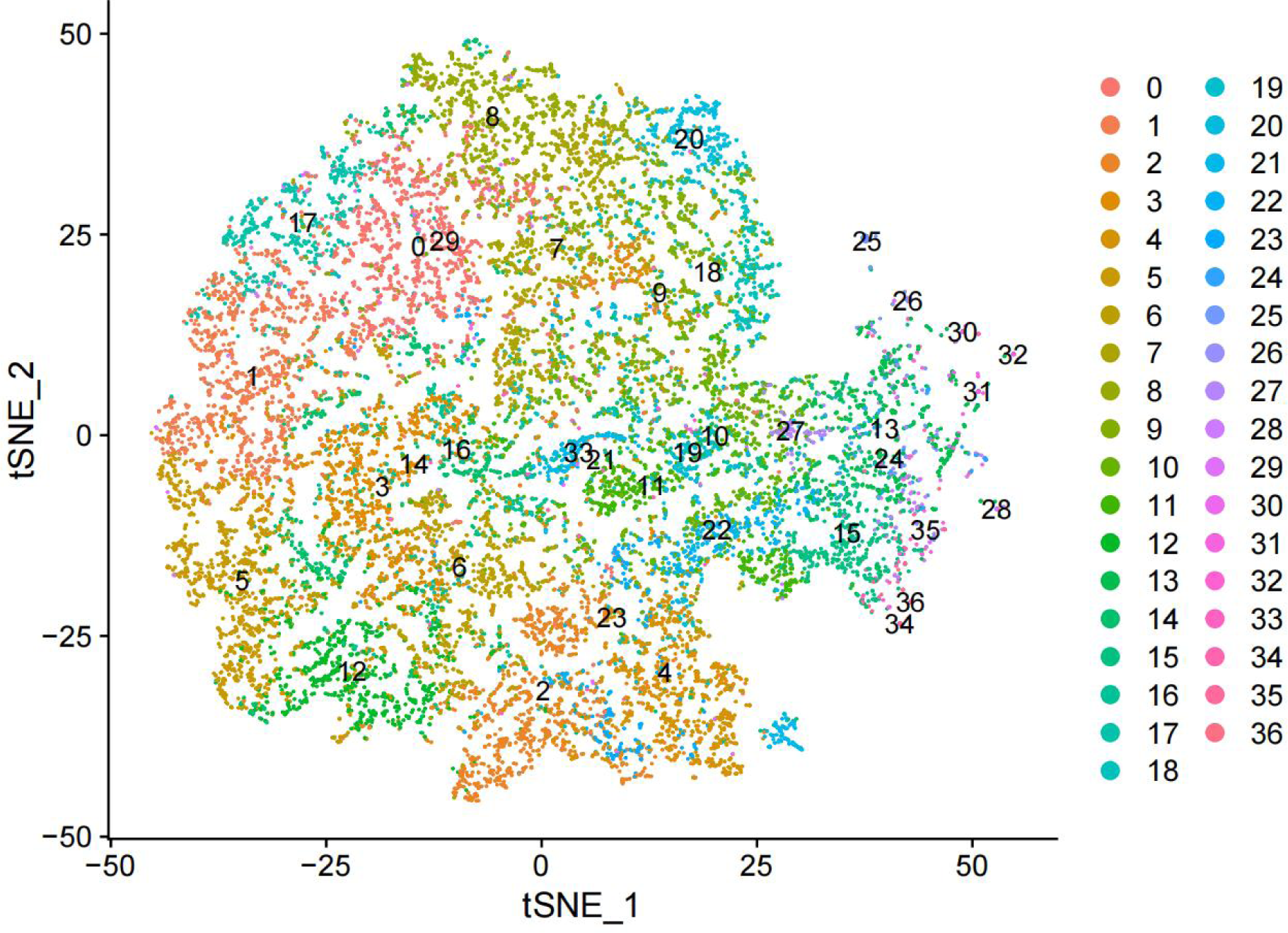
t-SNE visualization of gene clusters. Visualization of gene expression profiling clusters across NAFLD progression with the tdistributed stochastic neighbor embedding (t-SNE) statistical method. Each dot represents one protein coding gene (n = 17,946). A graph-based clustering approach was used. The dimensions of reduction were set to 1:20 and visualized with a resolution of 2.3.

**Supplementary Figure S2.**
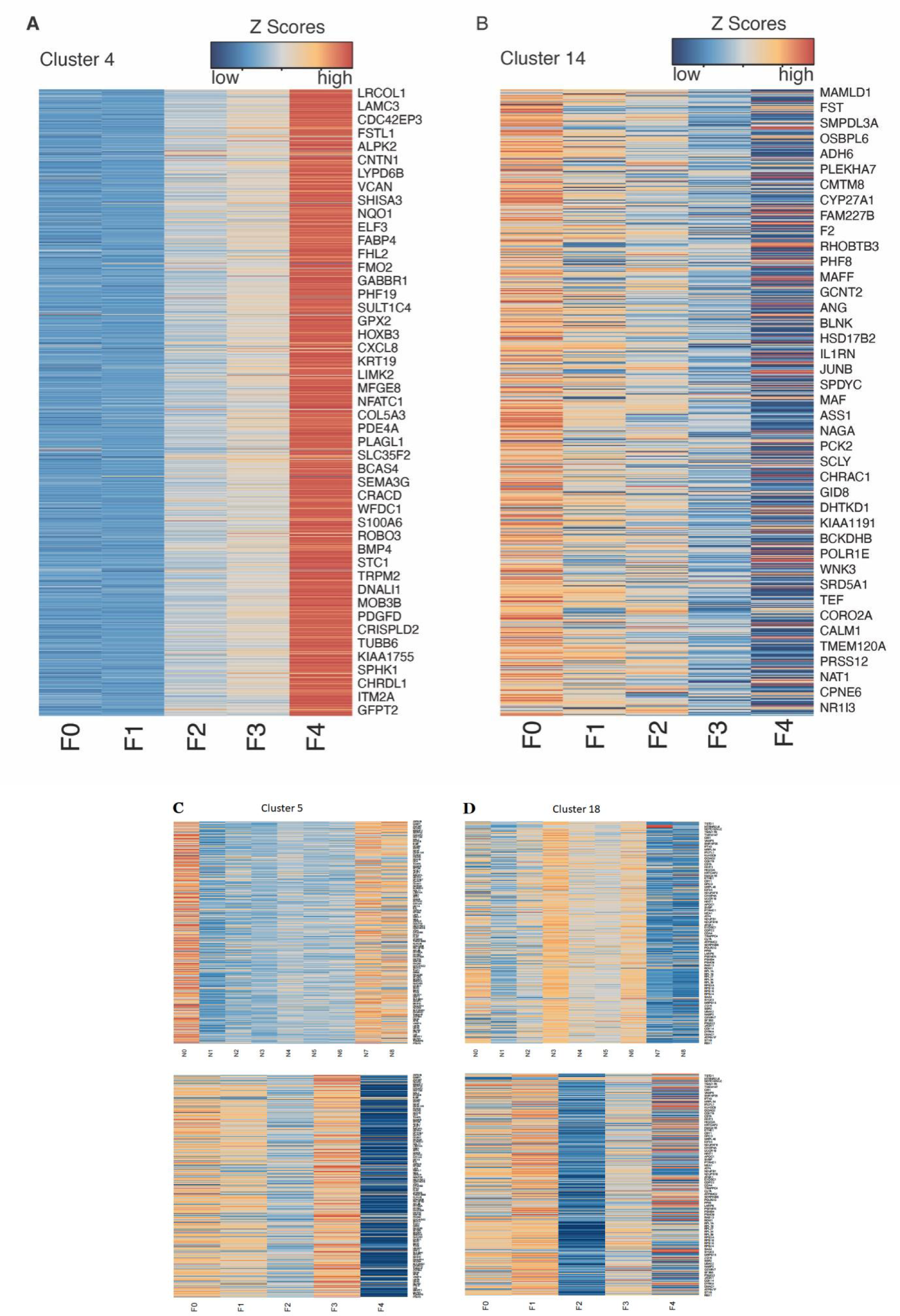
Heatmap presentation of gene expression profile along NAFLD progression. A. Heatmap presentation of 1021 up-regulated genes in cluster 4 associated with increasing fibrosis scores. B. Heatmap presentation of 643 down-regulated genes in cluster 14 associated with increasing fibrosis scores. C and D. Contrary to genes in cluster 4 and 14, C and D displayed gene clusters with chaotic gene expression patterns associated with both NAS scores and fibrosis scores.

**Supplementary Figure S3.**
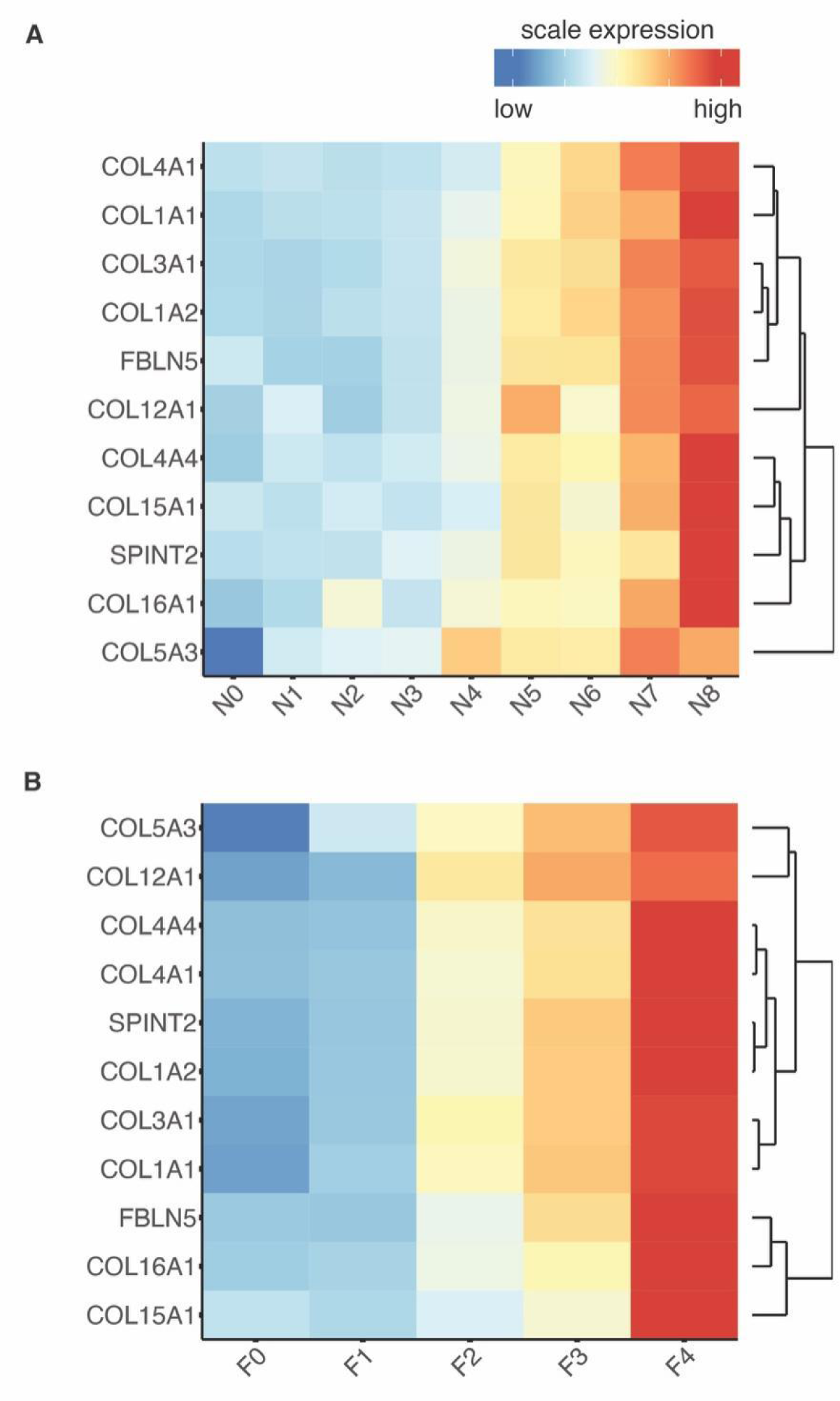
Heatmap presentation of ECM gene expression profile. A. Scale gene expression profile of multiple genes involved in the ECM process according to NAS scores. B. Scale gene expression profile of multiple genes involved in the ECM process according to fibrosis scores. Genes were clustered based on profile similarity. Genes expression level was scaled for heatmap presentation (also see the NAFLD-DB).

**Supplementary Figure S4.**
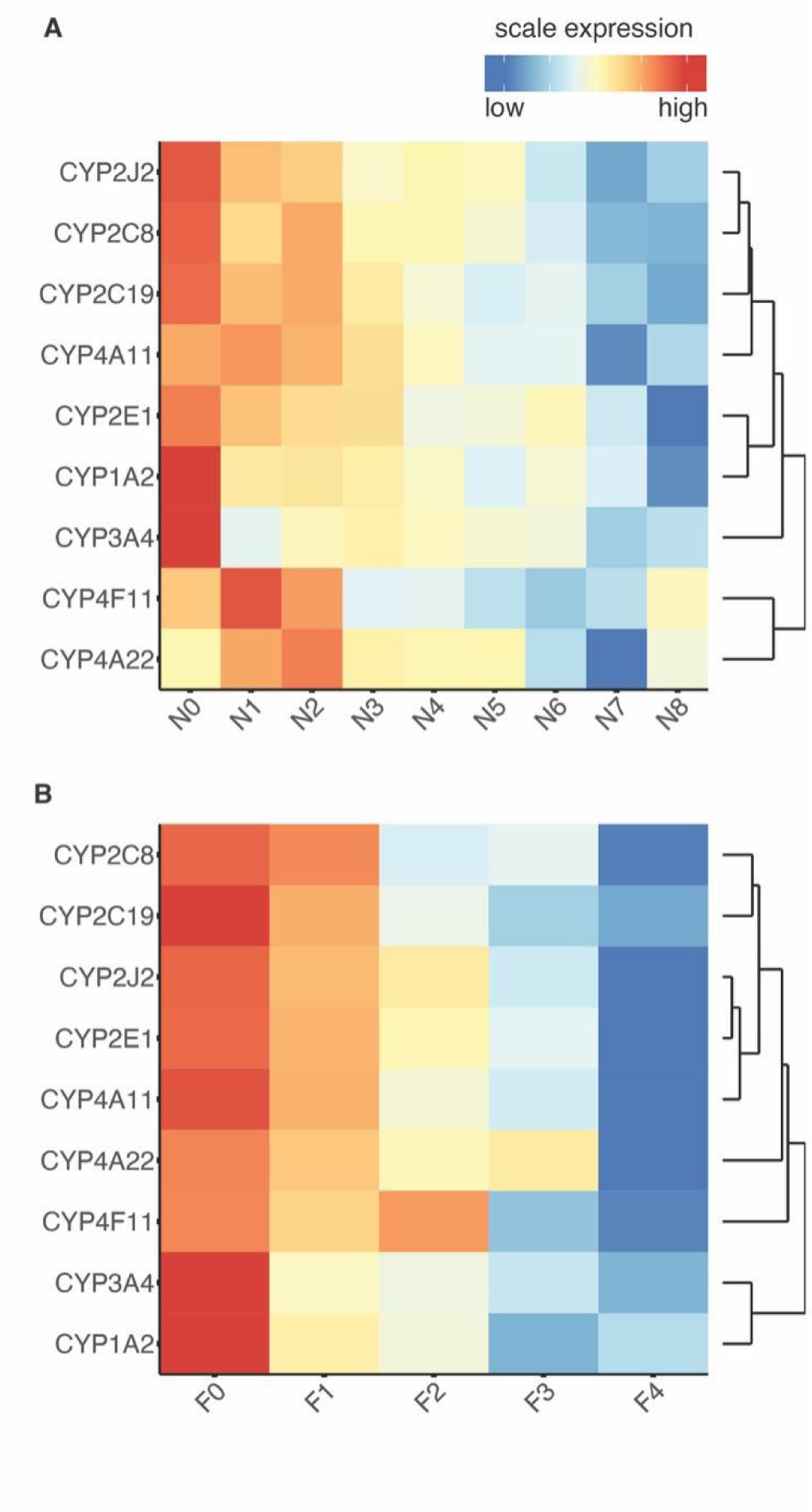
Heatmap presentation of Cytochrome P450 superfamily gene expression profile. A. Scale gene expression profile of multiple genes involved in the Cytochrome P450 superfamily according to NAS scores. B. Scale gene expression profile of multiple genes involved in the Cytochrome P450 superfamily according to fibrosis scores. Genes were clustered based on profile similarity. Genes expression level was scaled for heatmap presentation (also see the NAFLD-DB).

**Supplementary Figure S5.**
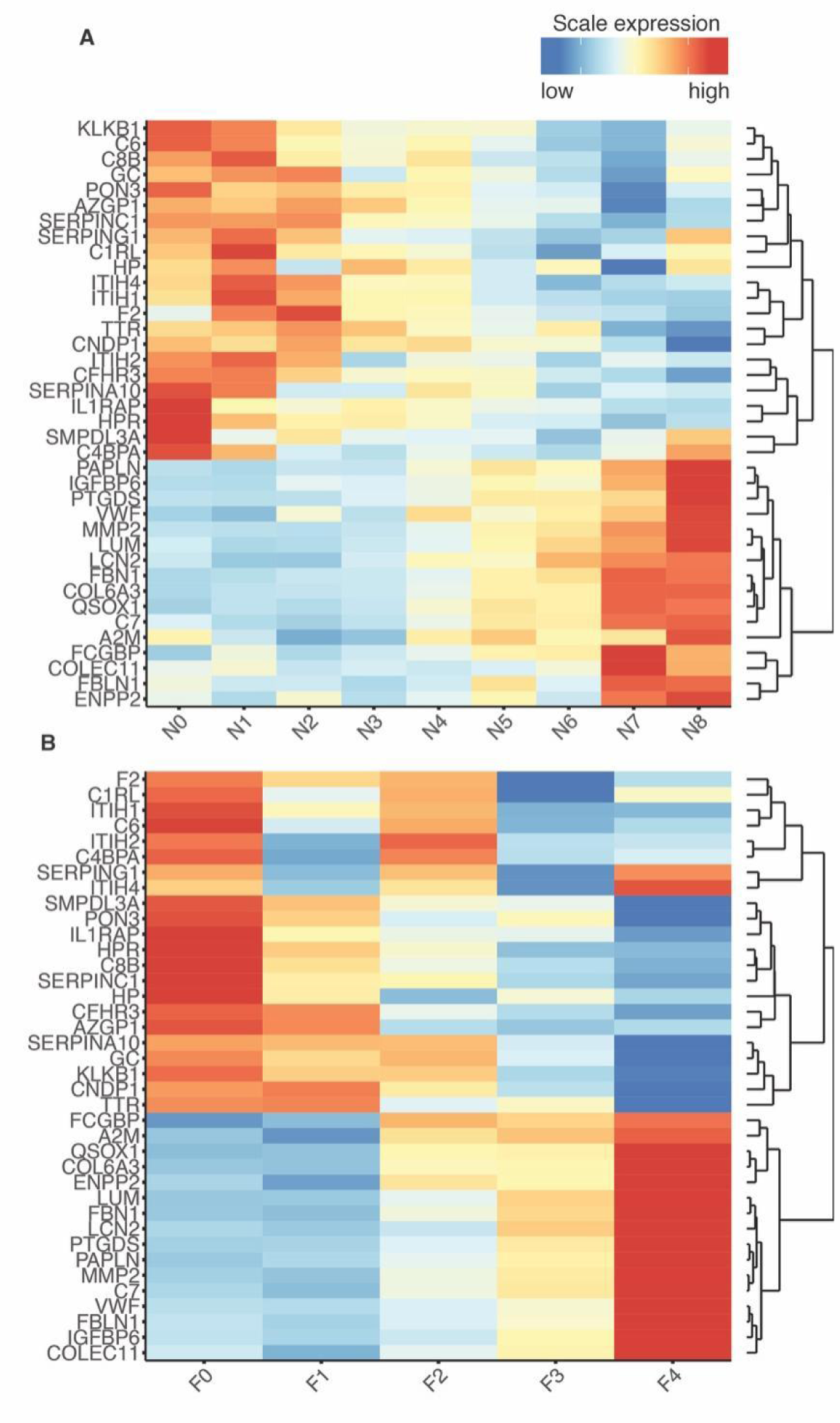
Heatmap presentation of expression profile for 32 biomarker genes. A. Scale gene expression profile of 38 biomarker genes (16 up-regulation, 22 downregulation) according to NAS scores. B. Scale gene expression profile of 38 biomarker genes according to fibrosis scores. Genes were clustered based on profile similarity. Genes expression level was scaled for heatmap presentation (also see the NAFLD-DB). This figure is related to Figure 2E.

## List of Supplementary Tables

**Supplementary Table 1**

The RNA-seq data of human liver samples.

**Supplementary Table 2**

GO enrichment results for genes in cluster 4 (up-regulation) and cluster 14 (down regulation).

**Supplementary Table 3**

List of secreting protein-encoding genes in cluster 4, and 14 of RNA-seq analysis. Representing PMID supporting that the candidate gene as potential biomarker for NAFLD was listed. Note: this is a noncomprehensive list.

**Supplementary Table 4**

Performance Characteristics of QSOX1, IL1RAP, and the QSOX1/IL1RAP ratio in proteomics data and the results of ELISA.

**Supplementary Table 5**

Metadata for NAFLD patients and control participants involved in the ELISA validation study.

